# Resting-state Functional Connectivity Predicts Cochlear-Implant Speech Outcomes

**DOI:** 10.1101/2024.01.30.24301908

**Authors:** Jamal Esmaelpoor, Tommy Peng, Beth Jelfs, Darren Mao, Maureen J. Shader, Colette M. McKay

## Abstract

**Background:** Cochlear implants (CIs) have revolutionized hearing restoration for individuals with severe or profound hearing loss. However, a substantial and unexplained variability persists in CI outcomes, even when considering subject-specific factors such as age and the duration of deafness. In this study, we explore the utility of resting-state functional near-infrared spectroscopy (fNIRS) recordings to predict speech understanding outcomes before and after CI implantation. Our hypothesis revolves around resting-state functional connectivity (FC) as a reflection of brain plasticity post-hearing loss and implantation. Specifically, we hypothesized that the average clustering coefficient in resting FC networks can capture this variation among CI users.

**Methods:** Twenty-two cochlear implant candidates participated in this study. Resting-state fNIRS data were collected pre-implantation and at one month, three months, and one year post-implantation. Speech understanding performance was assessed using CNC words in quiet and BKB sentences in noise one year post-implantation. Resting-state functional connectivity networks were constructed using regularized partial correlation, and the average clustering coefficient was measured in the signed weighted networks as a predictive measure for implantation outcomes.

**Results:** Our findings demonstrate a significant correlation between the average clustering coefficient in resting-state functional networks and speech understanding outcomes. Importantly, our analysis reveals that this measure provides unique information not accounted for by subject-specific factors such as age and duration of deafness.

**Conclusion:** This approach utilizes an easily deployable resting-state functional brain imaging metric to predict speech understanding outcomes in implant recipients. The results indicate that the average clustering coefficient, both pre and post implantation, correlates with speech understanding outcomes.

## 1. Introduction

Cochlear implants (CIs) have been instrumental in restoring hearing for individuals with severe to profound hearing loss. However, a significant proportion of CI users experience suboptimal outcomes or limited benefit from the implant [1]. While case-history factors such as age, duration of deafness, residual hearing, and previous experience with hearing aids have been investigated to explain this variability, they account for only a fraction of the variance in implantation outcomes [2; 3]. As a result, researchers are increasingly seeking more dependable predictors of speech understanding outcomes, particularly by examining changes in central cortical language networks. These networks are thought to be more closely linked to CI performance than the neural responses observed at lower levels of the auditory system, such as the brainstem or auditory nerves [4; 5; 6; 7].

Numerous studies have indicated neuroplastic changes in the brain following hearing loss and cochlear implantation, which can be broadly categorized as cross-modal and adaptive structural changes [6]. Sensory deprivation in one modality is known to lead to increased activity in brain regions associated with the remaining senses, resulting in the colonization of the primary cortical area by other modalities [8; 9]. Individuals with severe hearing loss often rely on intact senses, such as vision, to compensate for their hearing impairment. For instance, the auditory cortex in these individuals becomes more receptive to visual stimulation, enabling better visual localization and motion detection [10]. Some researchers argue that these cross-modal changes and the recruitment of the auditory cortex by other modalities may have maladaptive effects following cochlear implantation [9; 11]. For example, Doucet et al.[12] compared evoked potentials in response to visual stimuli of concentric gratings and found broader, anteriorly distributed cortical activations in patients with poorer speech understanding performance. Conversely, other studies suggest that these cross-modal changes in the auditory cortex may actually enhance CI users’ performance. They propose that the enhanced cross-modal plasticity in the auditory cortex improves visual speech understanding, leading to better lipreading abilities and speech understanding performance [13; 14; 7]. Anderson et al. [14] investigated cross-modal activation of the auditory cortex by visual speech before and 6 months after cochlear implantation, also based on fNIRS recordings. Their findings demonstrated a positive correlation between increased auditory cortex activation and CI recipients’ speech understanding ability 6 months after implantation. However, drawing definitive conclusions on the adaptive or maladaptive effects of cross-modal changes is challenging due to the wide variability in visual and audio stimuli employed in these studies. Neuroimaging evidence reveals that different types of speech stimuli elicit distinct brain activation patterns [15], including separate words and syllables, continuous speech for lingual visual and audio stimuli[16; 14; 17] and speech-like noise or checkerboards as non-lingual or visual stimuli [18; 7], each potentially leading to different activity patterns in the brain.

In addition to cross-modal changes, hearing loss is associated with significant adaptive structural changes in brain regions supporting auditory, language, and cognitive processing. These changes involve gray matter reduction in the inferior, middle, and superior temporal lobes, as well as the frontal and lingual gyrus. These structural alterations in the brain due to hearing loss affect auditory abilities and are linked to other cognitive dysfunctions, such as deficits in language function and semantic memory [19; 20; 21]. Furthermore, these structural changes can impact hearing abilities following cochlear implantation [22; 23]. Given that the brain’s structural connectivity network forms the basis for functional connectivity (FC) [24; 25; 26], assessing resting-state FC (also known as the intrinsic functional network) provides a means to evaluate these subtle changes in the brain network. Resting-state FC refers to the statistical dependence between activities in different brain regions (nodes in graph theory) in the absence of explicit stimuli or tasks. This approach has gained prominence in studying various brain disorders, including autism [27], Alzheimer’s [28], depression [29], and schizophrenia [30], as well as in investigating brain dynamics during learning [31] and aging [32]. The objective of this study was to identify a reliable indicator of cochlear implantation outcomes for individuals with postlingual deafness based on the resting-state functional networks of the brain.

This study aims to investigate the potential of resting-state FC as a reliable indicator of speech understanding outcomes in post-lingually deaf CI users. We propose that variations in the brain networks of these individuals can be reflected by the average clustering coefficient in the resting-state functional network, which quantifies the tendency of neighboring nodes to cluster together in graph theory [33]. To our knowledge, this is the first study to assess resting-state FC for evaluating brain plasticity in CI recipients and propose an indicator of speech understanding outcomes. By focusing on resting-state FC, our approach offers a streamlined, short, and straightforward assessment, avoiding the complexities associated with task-based experimental design and result interpretation. Furthermore, we utilized functional near-infrared spectroscopy (fNIRS) for brain imaging, a non-invasive and cost-effective optical imaging technology. Importantly, the implant device does not interfere with the fNIRS measurements [17].

## 2. Material and Methods

### 2.1. Participants

Twenty-seven adult CI recipients participated in the study. All participants were post-lingually deaf and implanted with Nucleus brand devices. Four of them did not complete the final test at 12 months. Besides, one subject did not get any benefit from the implantation after one year. So, they were removed from the study, and the results reported here are based on the data acquired from the remaining 22 subjects (mean age = 58.2±19.75, Table 1 provides the de-identified participant information.). The Human Research Ethics Committee of the Royal Victorian Eye and Ear Hospital approved this study (ethics approval 16.1262H), and all participants provided written informed consent.

**Table 1:** Demographic Information of Participants.

| Participant | Gender | Implant Ear | Deafness Duration(Year) | Implant Ear PTA* | Ear with Hearing Loss |
| --- | --- | --- | --- | --- | --- |
| Sub-1 | M | L | 2 | N.R. | L |
| Sub-2 | F | R | 10 | 100 | L & R |
| Sub-3 | M | L | 50 | 120 | L & R |
| Sub-4 | F | R | 20 | 112 | L & R |
| Sub-5 | F | L | 20 | 87 | L & R |
| Sub-6 | M | L | 5 | 82 | L |
| Sub-7 | F | L | 5 | 88 | L & R |
| Sub-8 | F | R | 1 | 57 | L & R |
| Sub-9 | M | R | 20 | 92 | L & R |
| Sub-10 | M | R | 12 | 107 | L & R |
| Sub-11 | M | L | 0.5 | 78 | L |
| Sub-12 | F | L | 18 | 70 | L & R |
| Sub-13 | M | L | 0.25 | 105 | L & R |
| Sub-14 | M | L | 20 | 114 | L & R |
| Sub-15 | M | L | 0.6 | 83 | L & R |
| Sub-16 | F | L | 0.75 | 90 | L |
| Sub-17 | F | L | 4 | 98 | L & R |
| Sub-18 | F | L | 46 | 100 | L & R |
| Sub-19 | F | L | 2 | 95 | L & R |
| Sub-20 | M | R | 13 | 103 | L & R |
| Sub-21 | M | L | 1.25 | 120 | L |
| Sub-22 | F | L | 20 | 120 | L & R |

\*PTA (dBHL) = pure-tone average of .5, 1, and 2 kHz hearing thresholds

Before implantation, the cognitive skills of our participants were assessed by doing trial-making tests A and B as described in [34], which were drawing lines between ascending numbers (test A) or drawing lines between numbers and letters in ascending order alternatively (test B). The participants were asked to complete the tasks as quickly and accurately as possible without removing the pencil from the paper. All of them had normal performance and passed the test criteria.

This study aimed to introduce predictors for speech understanding outcomes of CI users after one year. Therefore, the speech understanding outcomes were measured at 12 months post-implant. Each participant underwent two audio-only speech tests to measure their speech understanding performance. Direct audio inputs were applied to remove the effect of any residual hearing on the speech perception measurements with the device. The first test included 50 consonant-nucleus-consonant (CNC) words in quiet [35]. For this test, the overall scores were presented in percentages for both correct phonemes and words. The second test comprised 16 Bamford-Kowal-Bench Sentence (BKB) sentences in a multi-talker bubble noise environment [36]. The test’s score was determined based on the signal-to-noise ratio (SNR) needed to achieve 50% accuracy in word recognition. This SNR was dynamically adjusted during the test based on the participant’s response accuracy. If the accuracy rate exceeded 50%, the SNR was reduced by one unit, while it was increased by one unit if the accuracy rate fell below 50%. This process was repeated ten times, and the SNR values were averaged across the turning points of these iterations. Therefore, lower scores, indicating a lower SNR for achieving 50% word recognition, reflected better performance on the test. The SNR adjustment procedure for the BKB sentences began at 20 dB SNR. In both speech understanding tests, the speech was presented at a level of 65 dBA in the sound field, and the noise level was adapted to manipulate the SNR.

### 2.2. Data Acquisition

We recorded 5-minute resting-state fNIRS at four time points. The first recording was before implantation; the rest were at one, three, and 12 months post-switch on. The measurements were carried out using the NIRscout system (manufactured by NIRx company). The system uses LEDs with dual near-infrared wavelengths of 760 and 850 nm. Our montage included 16 sources and 16 detectors mounted on a 10-20 system cap which together built 54 channels. Two channels were short (5 mm distance between source and detector pairs) and used in the preprocessing steps to remove systemic artifacts from long channels [37]. Figure 1 shows the channel placements in the montage. Our regions of interest included the left and right auditory cortices and the left visual cortex.

**Figure 1:**
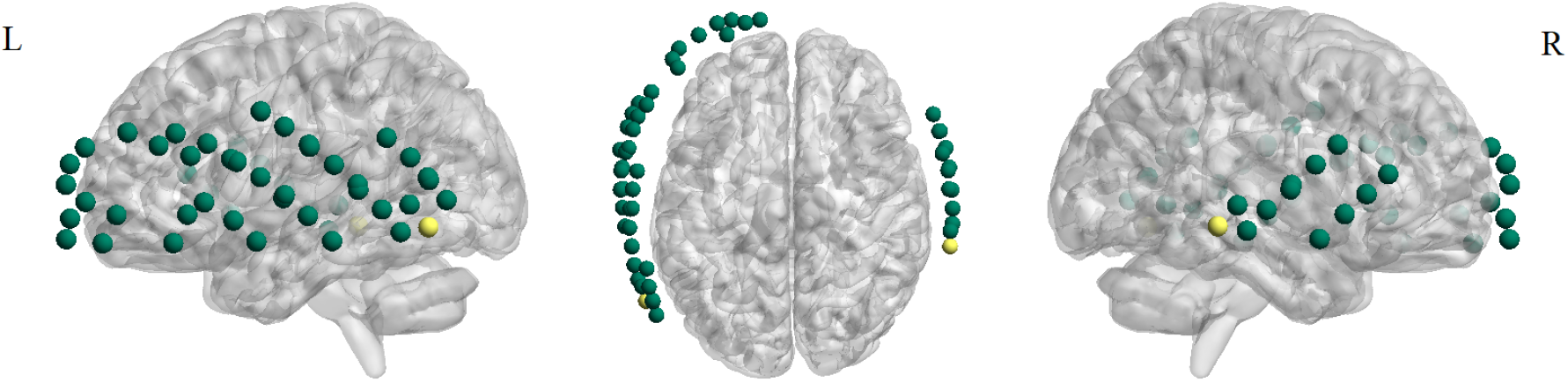
The montage we used for resting-state fNIRS recording. It included 52 long channels (green) and two short channels (yellow). The montage covered the right and left auditory cortices and the left visual cortex.

### 2.3. Data Preprocessing

We performed data preprocessing using the NIRS toolbox [38]. First, the raw recordings were converted to optical density. We evaluated the quality of channels using the scalp coupling index (SCI) [39]. Channels with SCI lower than 0.5 were indicated as bad ones and removed from the recordings. If the number of bad channels in each recording exceeded 26, the recording was removed from further analysis. Then, we applied the temporal derivative distribution repair (TDDR) method on the remained channels to improve signal quality by removing motion artifacts [40]. Afterward, the optical signals were converted to oxy- and de-oxyhemoglobin (HbO and HbR) based on the modified Beer-Lambert Law. Since long channels capture both cerebral activities and systemic artifacts, such as heartbeats, respiration, and Myer waves, we took two successive steps to mitigate the effect of these systemic artifacts and other noises. We used short channels for short-channel correction and applied a band-pass filter with a 0.02-0.40 Hz pass band. Short-channel correction regressed out short-channel signals that include systemic artifacts and no cerebral component [41]. The band-pass filter was applied to remove low-frequency artifacts like Myer waves or baseline drifts and high-frequency artifacts like heartbeats.

### 2.4. Functional Connectivity Matrix and Graph Construction

The connectivity analysis was conducted on data in the HbO format, as previous studies have demonstrated that it produces more robust coherence patterns and connectivity compared to HbR [42]. We measured FC between pairs of channels using regularized partial correlation. To illustrate constructing the functional brain network based on regularized partial correlation, suppose one performs multiple regression for each signal based on other signals as regressors. In that case, the regression slope for each regressor is proportional to the partial correlation coefficient between the signal and the regressor.

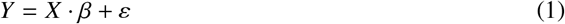

where X is an *(n*×*p)* matrix and includes *p* controlling regressors with n samples each. *β* is a *(p*×*1)* vector of regression slopes for each controlling variable. *ε* is a vector of n elements showing the error terms in the linear estimations of the *Y* variable. The ordinary least squares method to solve the equation leads to the highest possible dimension for *β* that is *p*. However, partial correlation is usually calculated using regularization techniques. Regularization applies extra penalties for network complexity. Doing so removes links between nodes (also known as edges in graph theory) that are likely to be spurious and help effectively to retrieve actual network structure [43; 26].

This study used regularized partial correlation based on L2-norm Ridge Regression (aka Tikhonov) to estimate FC networks [44]. The approach adds a term based on the squared sum of *β* values to the cost function:

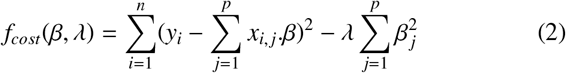

*λ* parameter that ranges from 0 to 1 penalizes the *β* weights and control density in the FC graph to get rid of spurious links in the network. Higher values result in further shrinkage of the edge weights and sparser networks.

In this study, we used the leave-one-out cross-validation method to optimize *λ* for each speech understanding test [45]. In this type of iterative method, one recording is left for testing each time, and others are used for training. The optimum *λ* value for each speech understanding test was chosen when the estimation error was minimum. Although leave-one-out is computationally expensive for large networks (e.g., in most fMRI studies), here, the cost was not a concern since our networks were relatively small (52 nodes).

### 2.5. Clustering Coefficient

In this study, the average clustering coefficients in the resting-state networks were considered features that represent CI users’ brain plasticity. We constructed signed weighted FC matrices based on partial correlation to measure connectivity between channels. Since correlation estimates for two signals are typically based on relatively short recordings, small amplitude correlations exist in the network as unstable estimates of correlations between nodes. Since they are distributed equally between positive and negative weights, their effects are expected to be canceled out when averaging node clustering coefficients in the network. Therefore, signed networks are more resistant to spurious connectetions than unsigned networks [46]. In the FC networks (with weights from -1 to 1), the average clustering coefficient represented the overall connection density in the areas covered by the montage. We measured the clustering coefficient of node *i* in the signed weighted networks as,

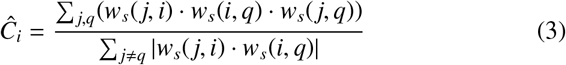

*w*_*s*_(*i, q*) is the edge weight connecting node *a* to node *b*. (*i, j, q*)s include all triangles with *i* as a vertex. The numerator is the sum of edge products of all triangles that include node *i* and the denominator equals the sum of absolute indirect traces between pairs of nodes that pass through node *i* [47; 46].

## 3. Results

### 3.1. Construction of Brain Functional Networks Based on Regularized Partial Correlation

We calculated the resting-state functional connectivity networks of the fNIRS recordings using regularized partial correlation [48]. Regularization applies extra penalties on network complexity to eliminate spurious connections, resulting in the shrinkage of the networks’ weight distributions. Figure 2 presents the effect of regularization on the functional connectivity matrix and weight distribution of a sample subject (subject seven, session three). As the figure shows, regularization has decreased the overall weight amplitudes and made the network sparser than the unregularized network by trying to remove spurious connections. Table 2 shows optimum regularization values (*λ*) for different tests across the sessions. The small (or zero) optimal *λ* values show that the regularization step had little (or no effect) on the complexity of networks in different sessions.

**Table 2:** Partial Correlation Regularization Parameter (*λ*) Values for Each Test Across the Sessions.

|  | Speech Tests |  |  |
| --- | --- | --- | --- |
|  | CNC words | CNC phonemes | BKB STR |
| Session 1 | 0 | 0 | 1.80E-5 |
| Session 2 | 0 | 0 | 0 |
| Session 3 | 1.32E-4 | 0 | 3.12E-4 |
| Session 4 | 3.80E-5 | 1.80E-5 | 9.40E-5 |

**Figure 2:**
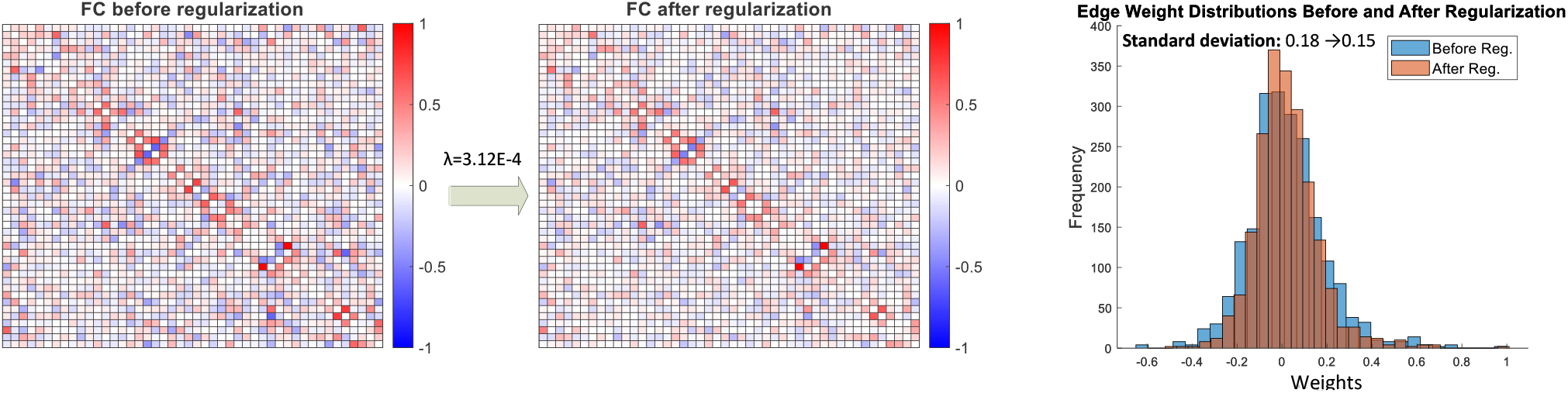
Regularization effect on the functional connectivity weights for a sample subject (subject seven, session three). Regularization applies extra penalties on network complexity to eliminate spurious connections, resulting in weight distribution shrinkage.

### 3.2. Clustering Coefficient at Resting State Correlates with Speech Understanding Scores at 12 Months Post-Implantation

We investigated the association between the average clustering coefficients in the signed weighted networks derived from the fourth recording session (at 12 months) and the respective speech understanding outcomes of cochlear implant recipients after one year. Our analysis unveiled a statistically significant correlation between the average clustering coefficients and all categories of speech understanding scores at the 12-month assessment: including CNC words and phonemes in quiet, as well as BKB sentences in noise (Figure 3).

**Figure 3:**
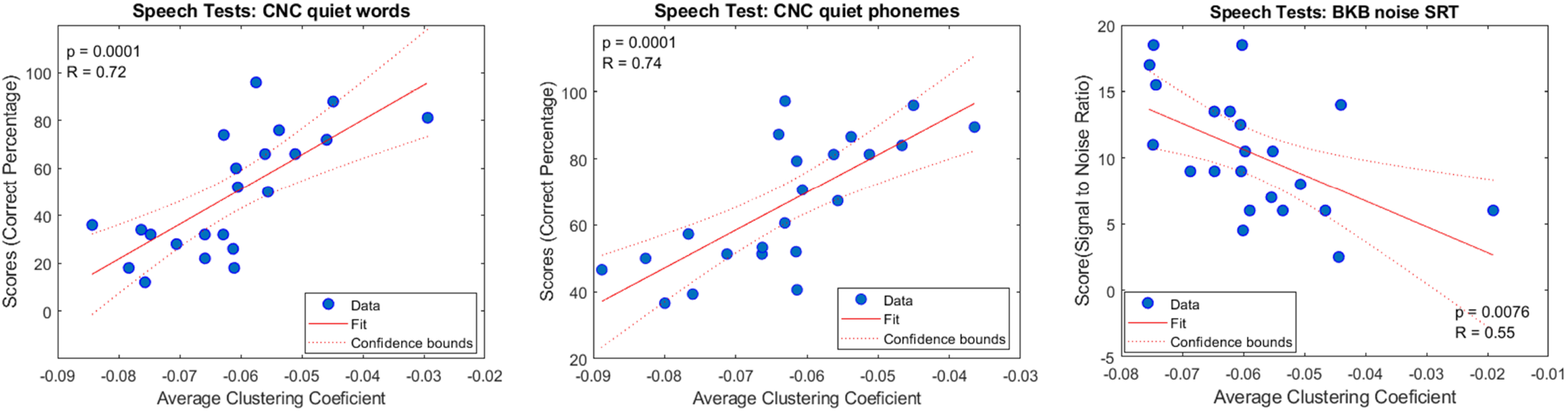
Average clustering coefficient at 12 months post-implantation correlates significantly with speech performance outcomes.

To ensure the robustness of our findings, we generated 30 null networks for each FC network following the method described by [49]. These null models were designed to serve as control graphs, maintaining connection weight distribution and node strength that closely resembled those of the primary networks. Figure 4 compares the p-values of the correlation between speech scores and the average clustering coefficient for the primary and null graphs. The figure shows that the correlation between the average clustering coefficient and test scores for the null models reduced considerably (higher p-values for null models).

**Figure 4:**
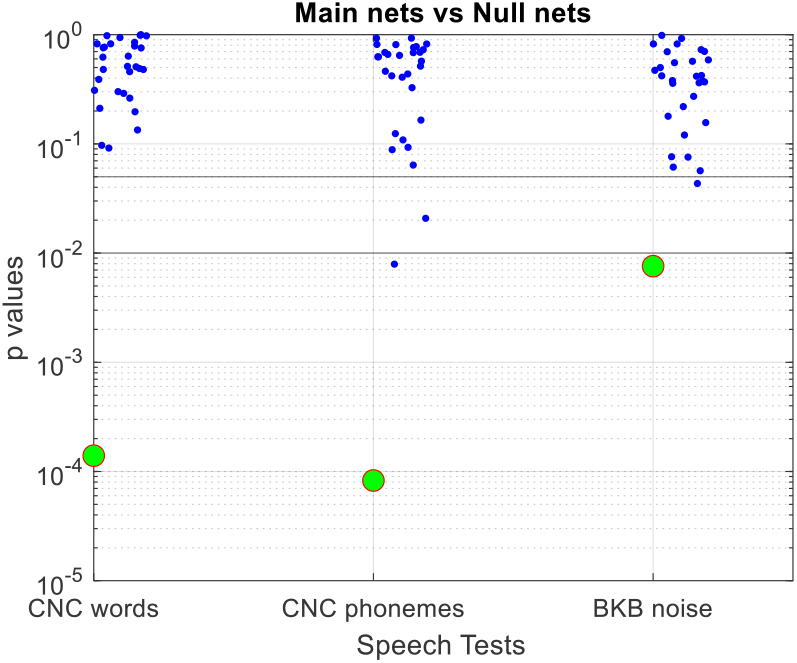
Comparing the significance of the correlations between the speech scores and the average clustering coefficient in the primary (green circles) and null graphs (blue dots). The figure shows that the correlation dropped for null models considerably.

### 3.3. Average Clustering Coefficient in Resting-State Functional Networks Predict Implantation Outcomes

We extended our investigation to assess the correlation between the average clustering coefficients within the fNIRS resting-state networks recorded before implantation, as well as at one month and three months post-implantation, and the corresponding speech scores obtained at the 12-month post-implantation assessment. This analysis was conducted to assess the predictive capabilities of the average clustering coefficient in the context of cochlear implantation outcomes. Figure 5 summarizes the correlation values (*R*) between average clustering coefficients in the resting-state networks of each session with speech understanding scores. As the results indicated, in many cases, the average clustering coefficients of the brain networks at different time points before one year were highly correlated with behavioral CI outcomes at 12 months post-implantation.

**Figure 5:**
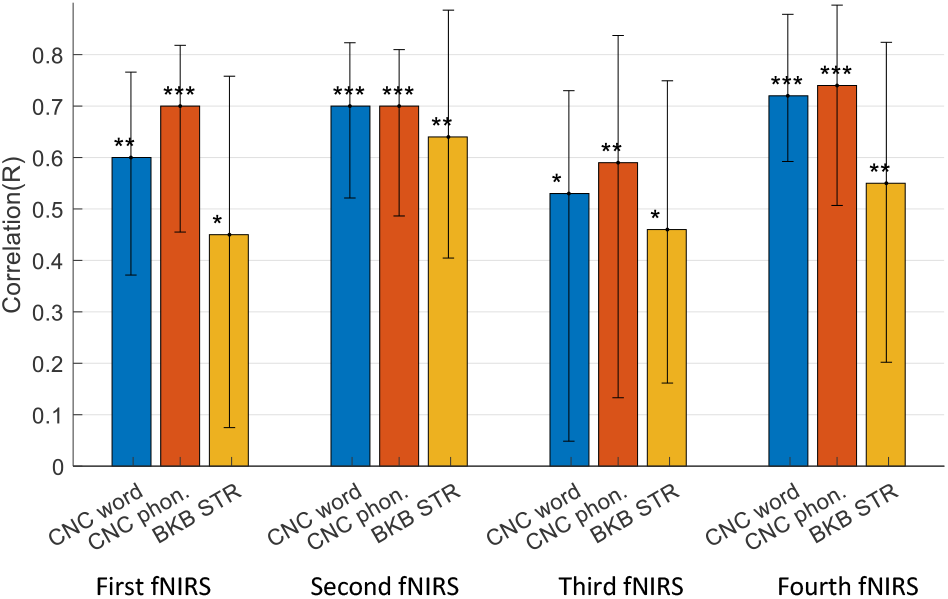
Correlation (*R*) between speech scores and their predictions based on the linear regressors in each fNIRS recording session. The error bars show the 95% confidence interval for 100 bootstrapped samples. The star marks (*), (**), and (***) indicate p-values smaller than the significance levels of *α* = 0.05, 0.01, and *α* = 0.001, respectively.

### 3.4. Average Clustering Coefficient Reveals Brain Plasticity

We conducted an analysis to investigate alterations in the average clustering coefficient within the brain networks of our subjects across various sessions. The findings underscored a significant increase in the average clustering coefficients during the three and twelve months post-implantation when compared to the baseline values recorded pre-implantation. However, no statistically significant changes were observed in the average clustering coefficient one month after the surgery (Figure 6).

**Figure 6:**
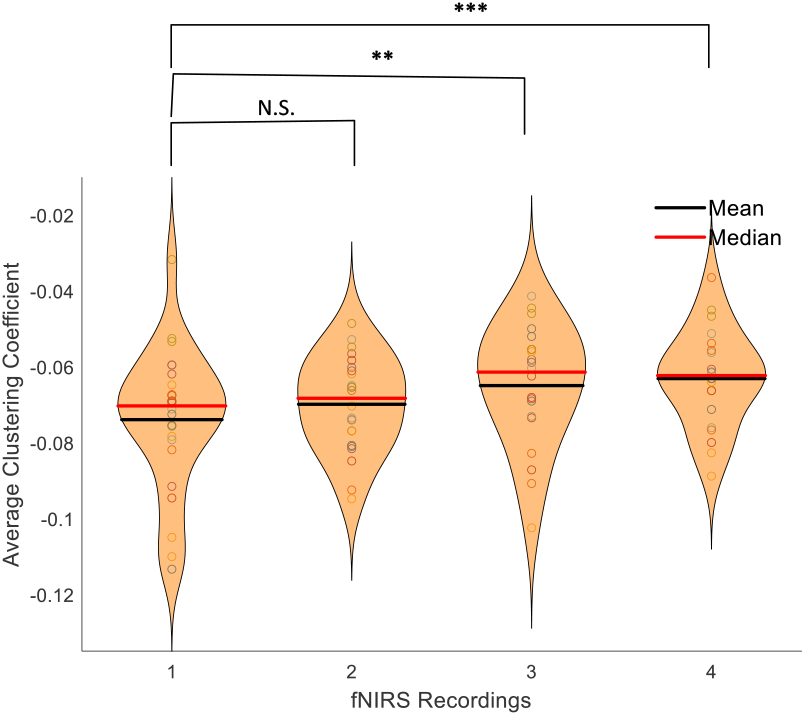
The distribution of the average clustering coefficients of our subjects in different sessions. The changes in average clustering coefficients were significant at three and 12 months but not in one month post-implantation. (N.S. stands for not significant.)

### 3.5. Average Clustering Coefficient Convey Unique Information Beyond Age and Deafness Duration

Age and deafness duration are critical subject history factors known to impact cochlear implantation outcomes. Typically, older individuals and those with a longer duration of deafness tend to exhibit poorer speech outcomes. As illustrated in Figure 7, the bar chart displays the correlation of these two factors with speech scores. Notably, the correlation of age with speech scores, specifically CNC words and phonemes, is relatively high and comparable to the correlation between the average clustering coefficient and these scores.

**Figure 7:**
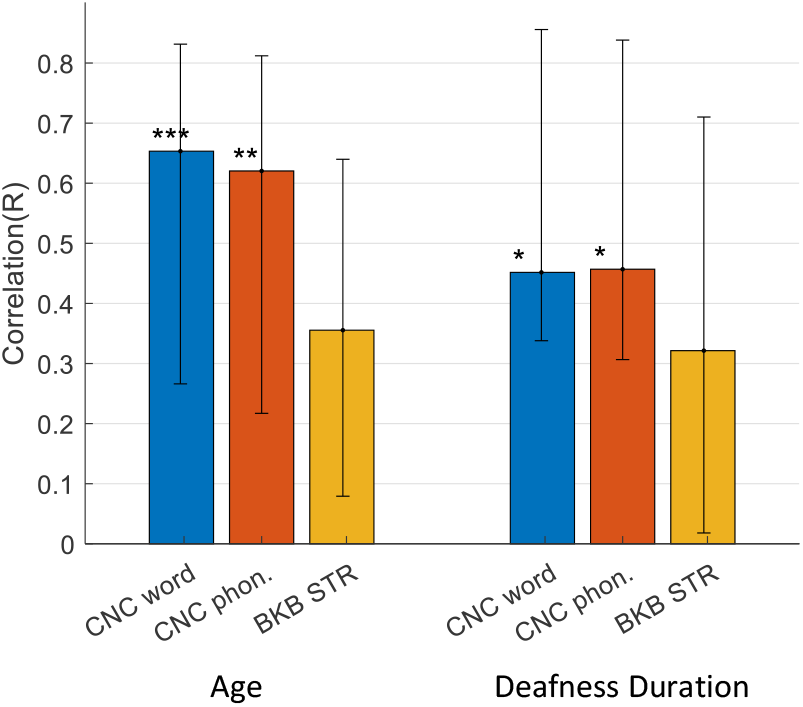
The correlation between age and deafness duration with speech performance outcomes.

To comprehensively evaluate the individual contributions of these factors, we conducted multivariable linear regression analyses for each session. Before conducting these tests, we carefully examined multicollinearity among the variables. Detecting multicollinearity is crucial because, while it does not reduce the explanatory power of the model, it can diminish the statistical significance of the independent variables. To evaluate multicollinearity, we calculated the variance inflation factor (VIF), which measures the intercorrelation among independent variables in a multiple regression model[50]. Importantly, the VIF values for all variables in the multivariable regression tasks were below 2, with a maximum VIF of 1.75. These low VIF values indicate that multicollinearity is not a significant concern in our analysis.

Figure 8 presents a comparison of the correlation coefficients between age, deafness duration, and average clustering coefficient in four fNIRS recordings, considering them as independent variables, and speech understanding scores as dependent variables. The black squares in the bar plot denote the improvement in correlations compared to using only the average clustering coefficient as a predictive factor (Figure 5). Table 3 displays the p-values from the F-statistic test, indicating the significance level of a variable when considering the other terms in the model. Variables that remain significantly important in the model, even when accounting for other variables, are highlighted in red in the table.

**Table 3:** Significance level of a variable considering other terms in the multi-regression model.

|  | First fNIRS |  |  | Second fNIRS |  |  | Third fNIRS |  |  | Fourth fNIRS |  |  |
| --- | --- | --- | --- | --- | --- | --- | --- | --- | --- | --- | --- | --- |
|  | Score 1 | Score 2 | Score 3 | Score 1 | Score 2 | Score 3 | Score 1 | Score 2 | Score 3 | Score 1 | Score 2 | Score 3 |
| Clustering Coef. | 0.118 | 0.020 | 0.241 | 0.029 | 0.028 | 0.029 | 0.341 | 0.084 | 0.207 | 0.002 | 0.002 | 0.032 |
| Age | 0.012 | 0.021 | 0.372 | 0.044 | 0.085 | 0.903 | 0.089 | 0.364 | 0.699 | 0.033 | 0.065 | 0.736 |
| Deafness Dur. | 0.719 | 0.907 | 0.864 | 0.613 | 0.564 | 0.937 | 0.260 | 0.120 | 0.518 | 0.208 | 0.228 | 0.334 |

**Figure 8:**
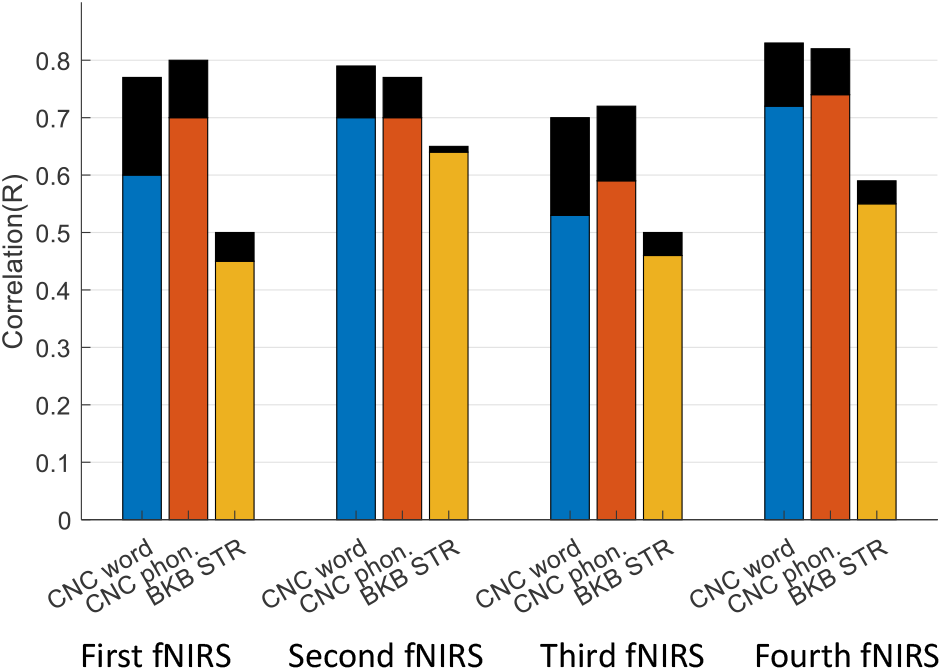
Correlations between age, deafness duration, and average clustering coefficient in four fNIRS recordings as independent variables, and speech understanding scores as dependent variables. Black squares indicate enhanced correlations compared to using the average clustering coefficient alone as a predictive factor.

### 3.6. Channel Density Influences the Correlation Between Average Clustering Coefficient and Implantation Outcomes

In our study, the computation of partial correlation between channel pairs takes into account the influence of other channels within the montage. This consideration is vital as it can impact the resulting correlation values and, consequently, the average clustering coefficient, which we utilize as a predictive measure for patients’ speech understanding performance following cochlear implantation. To delve deeper into this matter, we conducted experiments involving the removal of specific numbers of channels from the montage to assess the influence of channel density on our results. In each case, we randomly selected channels for removal from the setup and repeated this process 50 times for each number of omitted channels. Across different numbers of omitted channels, we calculated the average correlation between the average clustering coefficient and speech scores. Significantly, our results consistently indicate a trend: the reduction in channel density substantially decreased the accuracy of our proposed method across all sessions (refer to Figure 9 for an example, illustrating our findings based on the second fNIRS recording session).

**Figure 9:**
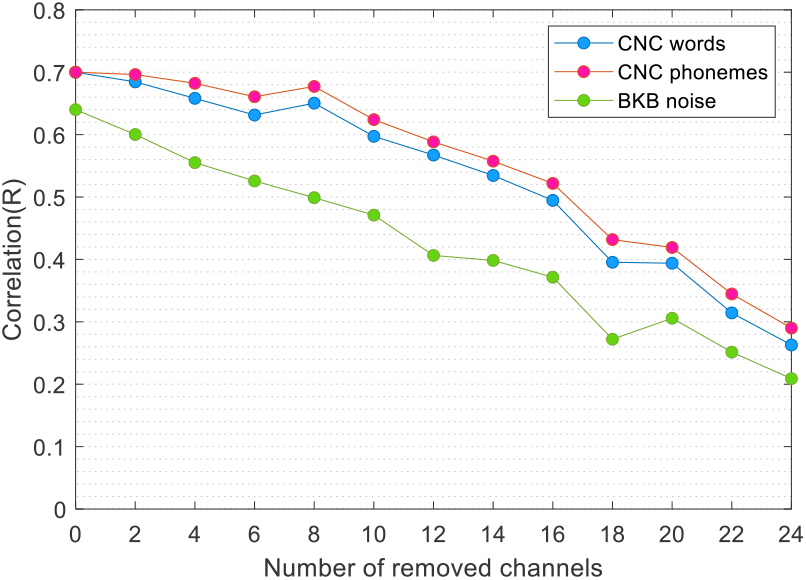
Decreasing the channel density by randomly removing specific numbers of channels from the setup results in a reduced algorithm precision, as indicated by the correlation between the average clustering coefficient and different speech scores. We conducted 50 iterations for each number of removed channels for the fourth session.

## 4. Discussion

The objective of the present study was to evaluate resting-state cortical activity using fNIRS and to investigate a reliable indicator of cochlear implantation outcomes. Previous studies on functional connectivity (FC) often centered around task-based experiments. However, there has been a shift towards resting-state FC studies, primarily for investigating brain abnormalities, group differences, and providing diagnostic and prognostic biomarkers. This shift is grounded in the fact that resting-state FC, also known as intrinsic brain FC, underpins task-based FC and operates across various brain states. The inconsistency in results from some task-based studies can be attributed to the diversity of tasks used in testing their hypotheses [7]. Therefore, using resting-state recordings simplifies the study of brain networks by eliminating the need for numerous task scenarios [51; 52]. Accordingly, this study aimed to propose, for the first time in the literature, a simple and reliable biomarker for speech understanding performance of CI users based on resting-state brain FC.

In this study, we used partial correlation to measure FC between pairs of fNIRS signals and create signed weighted functional networks. Partial correlation is effective in removing spurious connections and mitigating the effects of systemic noise, which is a significant concern in many fNIRS studies. Studies have shown that the FC networks created by regularized partial correlation correlate better with the underlying structural neural network compared to other connectivity measures such as Pearson correlation [26]. However, establishing the precise relationship between FC networks and structural networks, particularly for signed networks, is not straightforward, as most methods are developed for unsigned networks and do not consider the polarity of connections in the FC network [24; 53; 26].

The proposed measure in this study to interpret the variability in cochlear implantation outcomes was the average resting-state clustering coefficient in the language brain areas covered by our montage, including the right and left auditory cortices and the left visual cortex. The results demonstrated that this measure effectively explained the variation in cochlear implantation outcomes and exhibited high reproducibility across times pre- and post-implantation. Therefore, it can provide specialists with valuable insights before implantation to determine the potential benefits for the patient, guide adjustments to the device after implantation, or prescribe rehabilitation strategies to improve hearing performance.

The clustering coefficient of a node in a graph indicates how well its neighbors are connected or correlated with each other in a functional network. Our results showed a positive correlation between the average clustering coefficient and speech outcomes, indicating that subjects with higher average clustering coefficients in the areas covered by the montage had better speech understanding performances. These regions of interest included the left and right auditory cortices and the left visual cortex. Multiple studies have shown that hearing loss leads to brain atrophy in these brain areas involved in the language network, resulting in a reduction in neurons or neural connections [19; 20; 21]. The average clustering coefficient in the resting-state functional network can be interpreted as a measure of neural connection densities on a large scale since a high micro-scale correlation between resting-state functional and structural networks has been observed in many studies [24; 54]. Our study has shown that patients with higher average clustering coefficients (presumably interpreted as less atrophy) perform better with the CI.

The average clustering coefficient at different time points revealed gradual plasticity in the FC network after the CI switch-on (Figure 6). The results indicated that the average clustering coefficient increased after implantation due to the new stimuli provided by the device, aligning with the process of auditory recovery [4; 55]. Our findings showed that these changes were gradual and statistically significant after three months but insignificant for the first recordings taken one month after implantation. This pattern is in line with earlier research, which suggests that in the initial months following implantation, hearing remains suboptimal, and the sounds perceived are often challenging to decipher[56].

We also conducted a comparative analysis to assess the predictive performance of the average clustering coefficient in resting-state networks in relation to subject-specific historical factors such as age and deafness duration concerning speech outcomes. Deafness duration, as reported by subjects, is inherently subjective, leading to varying interpretations among different individuals. Consequently, it emerges as a less reliable predictor, and its correlation with performance outcomes in our dataset was notably weaker compared to the correlation with age. Our findings reveal that the inclusion of our proposed measure alongside subject-specific factors substantially enhances the accuracy of outcome estimation across all time steps, as illustrated in Figure 8. Moreover, the results highlight that our proposed measure consistently accounts for variability in speech understanding outcomes not elucidated by recipient history factors. This is especially notable during the second recording, one month post-implantation, a crucial period for clinicians fine-tuning the device (Table 3). This highlights the unique information about brain plasticity conveyed by our measure, information not captured by age or deafness duration alone.

The current study focused on the auditory and specific parts of the left visual cortex, limiting the assessment of brain plasticity. Including interconnected regions like motor and broader visual areas would provide a more comprehensive evaluation, capturing adaptive and cross-modal changes. Another metric, brain modularity, could also be explored. Modularity, representing densely connected subnetworks with sparse connections in real-life networks, underpins brain segregation [33; 57]. Studies suggest that sensory cortices, including hearing-related areas, experience atrophy and are influenced by remaining sensory areas like visual and motor regions [8; 58]. With hearing loss, the auditory cortex’s modularity may decrease due to reduced connections within the auditory module and increased connections with other areas, reflecting cross-modal plasticity. Post-implantation, the brain’s capacity to regain modularity might correlate with enhanced device performance.

The arrangement of channels in the montage plays a pivotal role in partial correlation calculations and, consequently, influences the average clustering coefficient utilized for post-implantation speech understanding predictions. Our experiments, involving random channel removals, clearly demonstrated that decreased channel density resulted in diminished algorithm precision. This underscores the critical significance of channel density in algorithm performance and suggests avenues for future research to optimize channel density, potentially enhancing the measure’s accuracy.

## 5. Conclusion and Future Works

This study aimed to interpret the significant variations in cochlear implantation outcomes among post-lingually deaf adults using resting-state functional connectivity. Our central goal was to devise an indicator illuminating the adaptive changes in the brain that occur after hearing loss and cochlear implantation. We harnessed fNIRS technology for brain imaging, with the advantage of not interfering with cochlear implant devices. Our chosen metric was the average clustering coefficient, calculated within specific brain regions: the left and right auditory cortices and the left visual cortex.

Our results consistently unveiled a correlation between the pre- and one-year post-implantation average clustering coefficient and speech understanding outcomes. Moreover, we observed a notable increase in the average clustering coefficient within the initial three months post-implantation, suggesting brain plasticity and the potential for auditory recovery. Crucially, our metric provided unique insights beyond conventional factors like age and deafness duration, significantly enhancing prediction accuracy when combined with these variables.

To deepen our comprehension of brain plasticity, future research might consider broadening the scope to encompass additional cortical regions. Including areas such as the motor cortex and other sections of the visual cortex, which undergo cross-modal adaptations as the auditory cortex post-hearing loss, could unveil a more comprehensive spectrum of brain plasticity. Thus, future studies should explore graph features to capture these diverse aspects effectively.

Furthermore, our study highlighted the pivotal role of channel density within the imaging setup in predicting cochlear implant outcomes. Consequently, we advocate for channel density to become a central consideration in future investigations, as it significantly influences the prediction accuracy of the proposed method.

## Data Availability

All data produced in the present study are available upon reasonable request to the authors

## Author Contributions

CM designed the study and contributed towards data acquisition. JE, TP, BJ, and DM performed data analysis. All authors interpreted the data. JE drafted the manuscript. All authors critically reviewed the manuscript before submission.

## Acknowledgement

This study was supported by a grant from the Passe and Williams Foundation to Colette M. McKay. Tommy Peng was supported by a Junior Fellowship from the Passe and Williams Foundation. Jamal Esmaelpoor was supported by the University of Melbourne Research Scholarship. The Bionics Institute acknowledges the support it receives from the Victorian Government through its Operational Infrastructure Support Program.

